# The Algerian chapter of SARS-CoV-2 pandemic: An evolutionary, genetic, and epidemiological prospect of the first wave

**DOI:** 10.1101/2020.11.19.20235135

**Authors:** Safia Zeghbib, Balázs Somogyi, Brigitta Zana, Gábor Kemenesi, Róbert Herczeg, Fawzi Derrar, Ferenc Jakab

## Abstract

To explore the SARS-CoV-2 early pandemic in Algeria, a dataset comprising forty-three genomes originating from SARS-CoV-2 sampled from Algeria and other countries worldwide, from 24 December 2019 through 8 March 2020, of which, were thoroughly examined. While performing a multi-component analysis regarding the Algerian outbreak, the toolkit of phylogenetic, phylodynamic, haplotype analyses and genomic analysis were effectively implemented. We estimated the TMRCA in reference to the Algerian pandemic and highlighted both the introduction of the disease originating in France and the missing data depicted in the transmission loop. Most importantly, we unveiled mutational patterns, recombination events and the relatedness regarding the Algerian sequences to the dataset. Our results revealed the unique amino-acid replacement L129F in the orf3a gene in Algeria_EPI_ISL_418241. Additionally, a connection between Algeria_EPI_ISL_420037 and sequences originating from the USA was observed through a USA characteristic amino-acid replacement T1004I in the *nsp3* gene, found in the aforementioned Algerian sequence. Lastly, we assessed the Algerian mitigation measures regarding disease containment using statistical analyses.

**Author summary:** A novel human coronavirus, specifically SARS-CoV-2, emerged in China in late 2019. In Algeria, the contact tracing revealed the introduction of the disease originated in France, however, the intricate dynamics regarding the disease remain unexplored. In this study, we attempt to portray our perspective regarding the evolutionary, genetic and epidemiological aspects of the early pandemic in Algeria during the spring of 2020, through the use of time scaled phylogeny, phylodynamic and mutational pattern characterization and exploration. Additionally, we assessed the efficiency of the implemented mitigation measures using statistical analysis. The results supported the virus introduction from France and highlighted an Algerian characteristic amino-acid replacement in addition to a relatedness to the USA sequences. Moreover, we revealed an indirect contamination among the three sampled patients. Therefore, our analysis is a starting point for further investigations and emphasize the importance regarding intensive sequencing and genome exploration for mitigation measures implementation, and both drug and vaccine development.

## Introduction

Historically, the emergence of SARS /Severe Acute Respiratory Syndrome dates back to 2002, and MERS/the Middle East Respiratory Syndrome epidemic erupted in 2012. In late 2019, the third highly pathogenic human coronavirus was first identified in Wuhan, China, as the epicenter and cause of a pneumonia outbreak [1]. The novel virus was aptly identified as a severe acute respiratory syndrome coronavirus 2 /SARS-CoV-2, and the primary cause of the coronavirus disease 19/COVID-19 [2]. Owing to travel, it subsequently spread worldwide and was declared a pandemic by the World Health Organization/WHO, and today is considered a major public health concern [3]. Similarly to MERS coronavirus and SARS coronavirus, the SARS-CoV-2 belongs to the *betacoronavirus* genus and Sarbecovirus subgenus, and related to SARS coronavirus with roughly 80% identity at the nucleotide level [1]. Moreover, it is likely bat originated, whereas the intermediate host if any, remains blurred, despite different hypotheses [4].

In Algeria, 25 February marks the first imported case and was registered in the southern portion of the country, in which an Italian employee tested positive. This was well contained, and no additional cases were reported until the beginning of March, which currently, is considered as the onset of an outbreak in Blida in Northern Algeria, as two more cases were recorded following a visiting family member from France [5]. In the present study, we aim to understand the dynamics associated with the transmission in reference to the first three diagnosed cases in Blida, of which, is the epicenter of the pandemic in Algeria and further characterize the identified Algerian SARS-CoV-2 genomes. Notably, this is of major importance regarding disease containment and both vaccine and drug development. Therefore, a dataset representing forty-one SARS-CoV-2 sequences comprising the three Algerian sequences deposited and freely available from the GISAID database were analyzed. To effectively manage the sequencing procedure, we employed the Beast v1.10.4 package for evolutionary, phylodynamic, and phylogeographic investigations, and, additionally, implemented POPART software to create a haplotype network analysis, to demonstrate introductions, local transmissions, and to further understand the disease spread and evolution. Additionally, both MEGA X and DnaSP v6.12.03 were used for further genome exploration. The results emphasized the absence of recombination within our dataset. We noted a similarity regarding the codon usage pattern across the Algerian sequences when compared with the reference sequence. Similarly, a mutational heterogeneity in both the nucleotide and protein levels across the Algerian genomes was clearly observed, highlighting both negative and positive selection. Additionally, the present outcome undeniably supports the disease introduction from France. However, it likely demonstrates a relationship to USA sequences and distinctly, highlights how missing unsampled data linking the three diagnosed infections were not a result of direct contamination among the three patients and implies undiagnosed infections during the early stage of the outbreak.

## Results

### Evolutionary phylogenetic, phylogeographic and phylodynamic analyses

The resultant log files from both the strict and the lognormal uncorrelated relaxed clock models were analyzed using the tracer. The posterior density regarding the lognormal standard deviation excludes zero indicating no rate variation. Thus, the strict clock model was rejected in favor of the relaxed clock model. The date of the most recent common ancestor (MRCA) of the global pandemic and its 95% highest posterior density interval (HPD) was 17/12/2019 [2019-12-08, 2019-12-23] under the exponential growth tree prior. Whereas, it was 16/12/2019 [2019-12-02, 2019-12-23] when using the Bayesian skyline plot model. Similarly, the estimated MRCA regarding the Algerian pandemic was 20/02/2020 [2020-02-13, 2020-02-27] and 21/02/2020 [2020-02-15, 2020-02-28] under the respective aforementioned tree priors. The SARS-CoV-2 estimated evolutionary rate was 0.001 [0.001 and 0.002] under both tree priors.

Furthermore, based on the maximum clade credibility tree (MCC) (Fig 1), the three Algerian sequences formed a monophyletic clade (Posterior probabilities: PP=1 and PP=0.99) and were closely related to sequences from France (PP=0.99). Thus, indicating the disease introduction originating from France and migrating to Algeria then spreading through local transmissions. However, the Algerian sequence hCoV-19/Algeria/G0860_2262/2020 was considered as an outgroup vis-à-vis hCoV-19/Algeria/G0640_2265/2020 and hCoV-19/Algeria/G0638_2264/2020) implying missing sequences.

**Fig 1.**
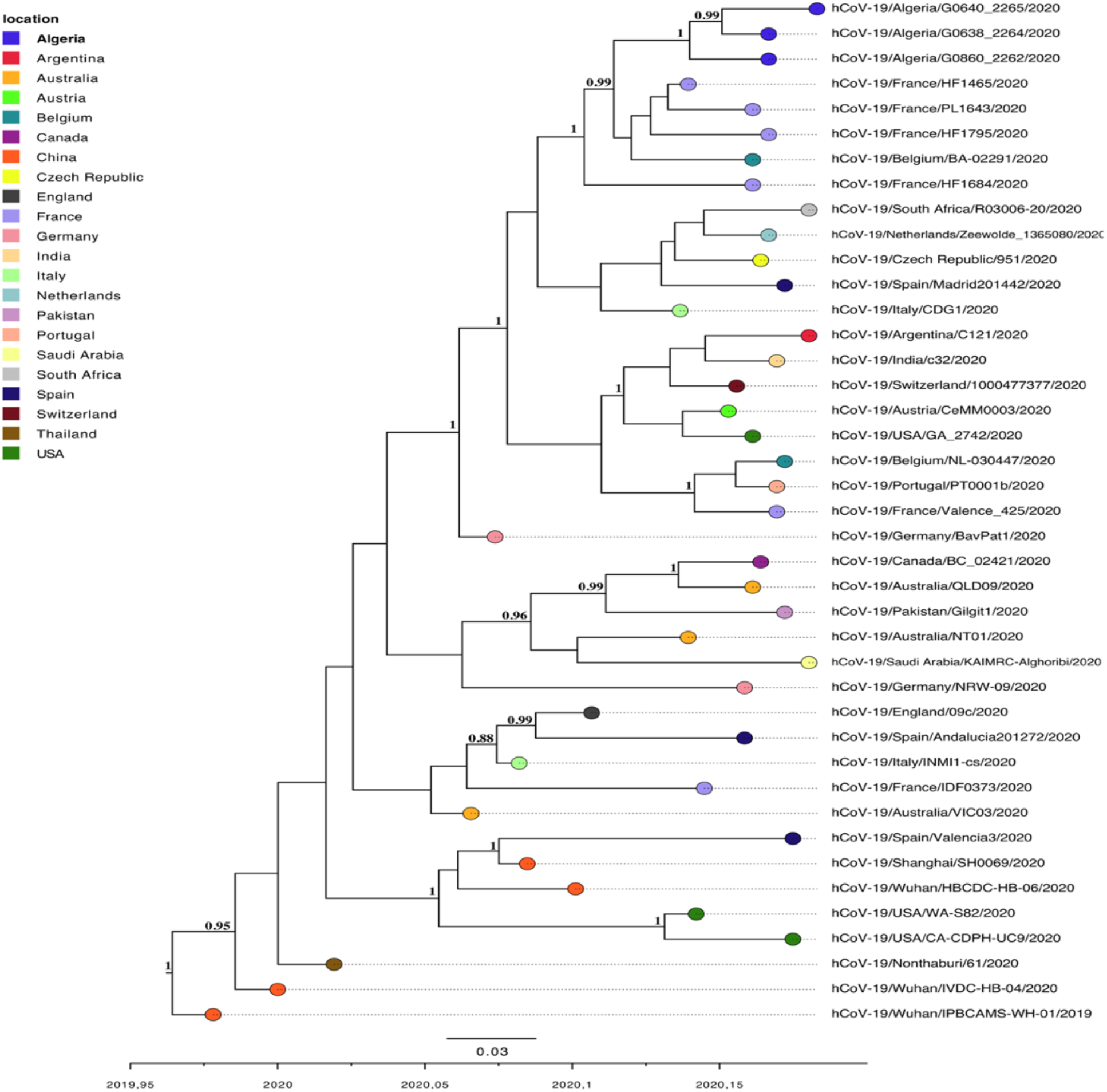
The maximum clade credibility tree was inferred based on forty-one SARS-CoV-2 full genomes. Branches are time scaled in decimal years. Only posterior probabilities ≥ 88 were shown on the tree. Labels on the right of the figure represent the sampling locations and are depicted as circles at the tips of the tree. This Bayesian phylogenetic tree was implemented using a relaxed molecular clock and a discrete phytogeographic approach using BEAST v1.10.4.

Likewise, phylogeographic analyses confirmed China as the origin of the pandemic (Fig 2A). Moreover, while relying on the Bayesian stochastic search variable selection /BSSVS and Bayes factor calculation, there was very strong supportive evidence mounting in support of the disease introduction from France to Algeria, substantiated by a Bayes factor = 42.04 (Figs 2B and 2C). Overall, the route depicting its global spread was first demonstrated to have originated in Asia then shortly thereafter, migrated to Europe (3<BF<6.11) and followed later to Africa (BF=42.04) (Fig 2D)

**Fig 2.**
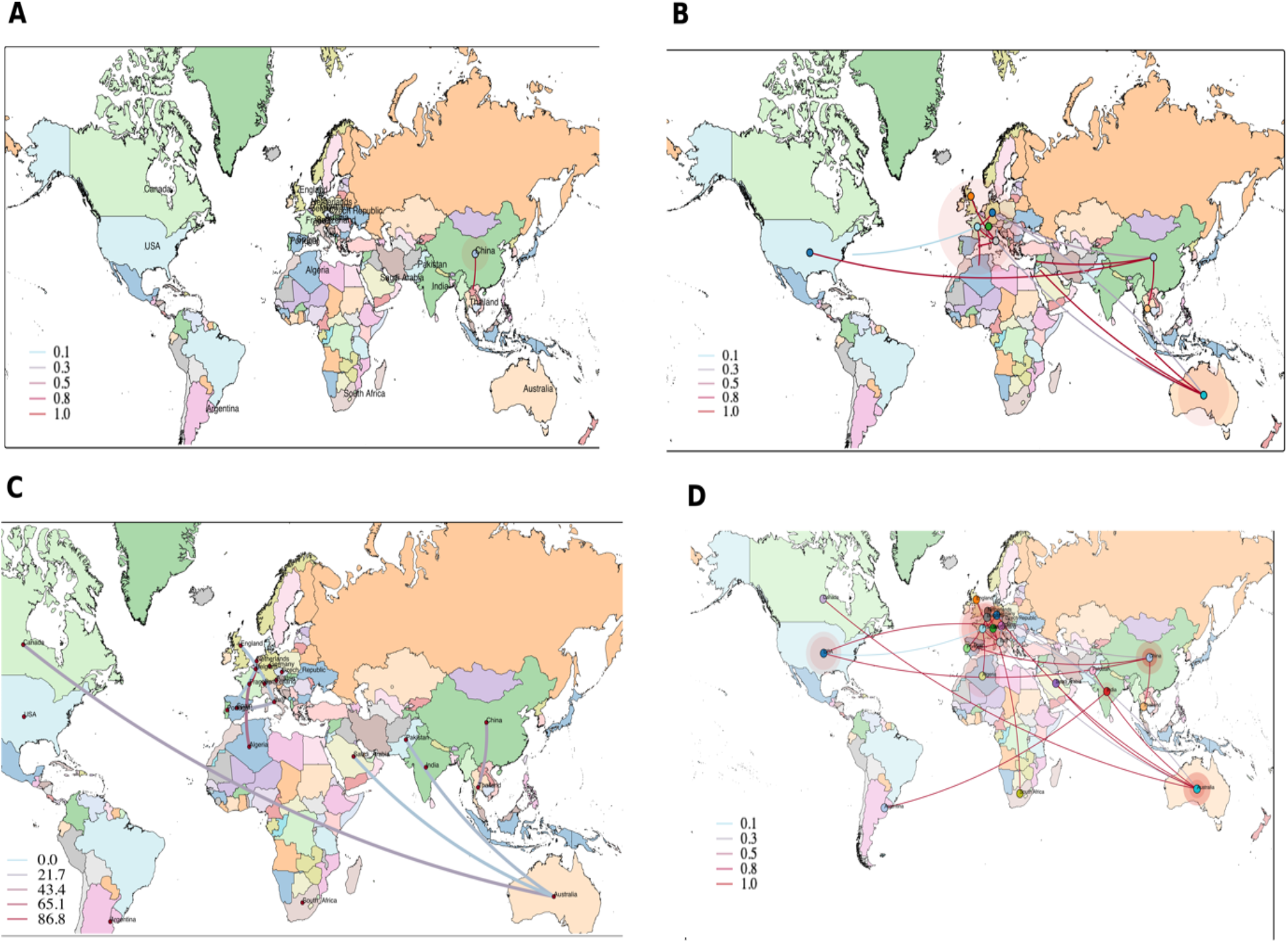
**(A)** China is the pandemic epicenter. **(B)** The introduction of SARS-CoV-2 originating from France to Algeria. **(C)** Bayes factor test regarding a discrete phylogeographic diffusion, only rates supported by a BF greater than ten are displayed. **(D)** The dispersal pattern regarding SARS-CoV-2 among different countries within the dataset. Lines between locations represent branches in the MCC tree. Branches are color coded according to their posterior probabilities in **A, B, D** and the Bayes factor in **C**. Blue-red color gradient represents a variation from the lowest to its highest value, respectively.

To obtain an overview regarding the demographic history within the dataset, we relayed to both parametric and non-parametric models, respectively. The exponential growth model demonstrated an increase in reference to the population size with a growth rate estimation of 0.06/day and a 95% HPD interval of [0.03, 0.09], thus the doubling time was predicted to be 11.7 days (Fig 3A). From another perspective, the skyline plot model displayed a logistic growth with a plateau phase beginning 15 February (Fig 3B).

**Fig 3.**
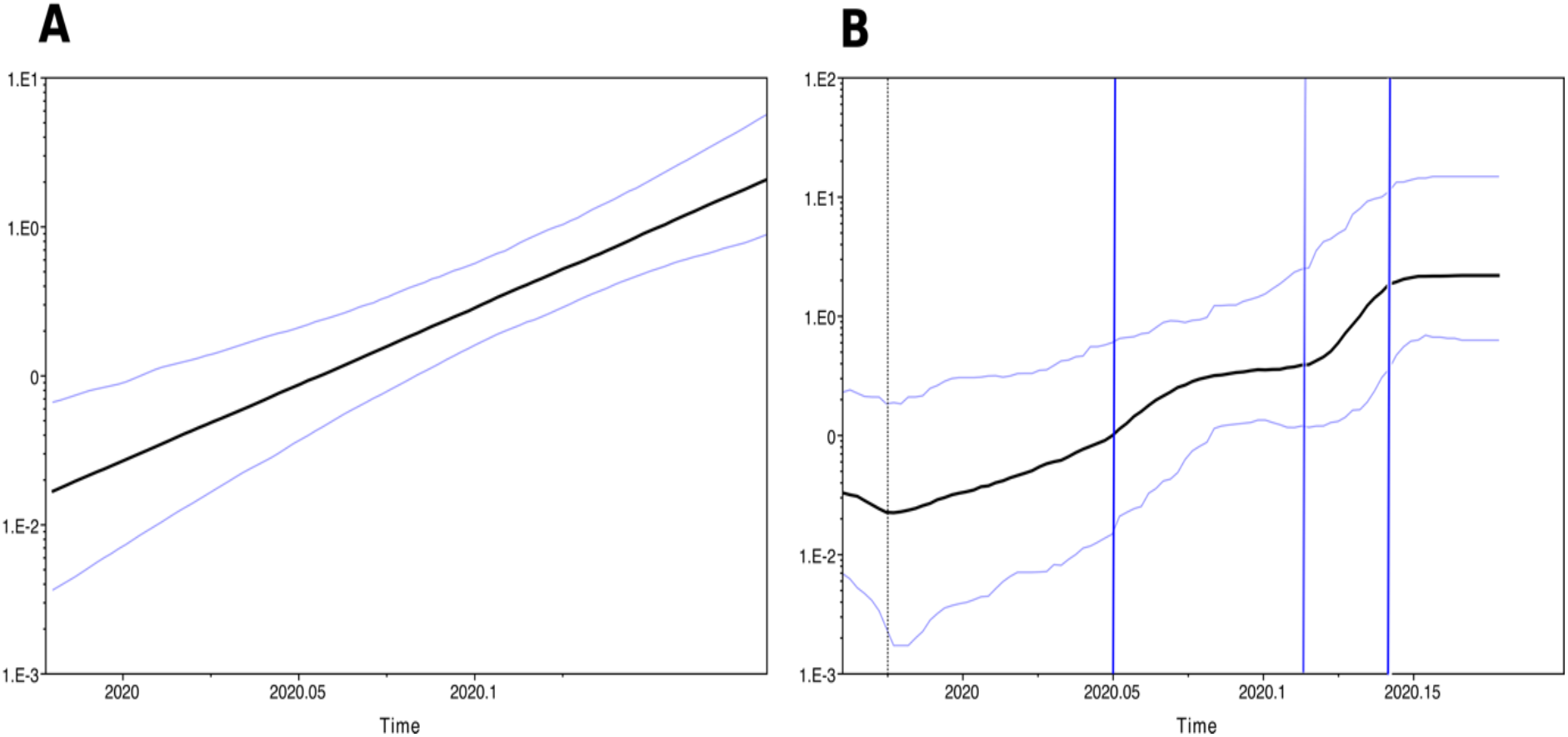
**(A)** Effective population size of SARS-CoV-2 and exponential growth rate. **(B)** Bayesian skyline plot of 41 SARS-CoV-2 genomes. The x-axis depicts the time scale in decimal years, and the y-axis symbolizes the logarithmic Neτ scale (Ne is the effective population size and τ is the generation time). The black solid line represents the median estimates and the cyan lines denote 95% highest posterior density. In panel **(B)**, the vertical blue lines highlight the different growth phases while the doted black line expresses the MRCA.

### Genome analyses

Based on p-distance pairwise calculations within and among the various groups defined by sequence locations, the Algerian sequences group exhibited a mean genetic distance of 0.01% (SE=0.0001). Notably, nucleotide divergence among sequences from different locations ranged between 0.02% and 0.05%. The lowest divergence was observed among the Algerian sequences and, also sequences originating from France, Czech Republic, Netherlands, and South Africa (SE=0.001). In contrast, the Algerian sequences and sequences originating from the USA, Canada, and Saudi Arabia exhibited the highest divergence (0.05%, SE=0.0001) (Table S1).

Moreover, codon usage pattern analysis pertaining to the compositional characteristics in the Reference sequence and the Algerian sequences demonstrated the value of overall G+C content in *ORF1b* and *S* genes is 37%, while in *ORF1*a and *E* genes is 38%, the higher values are observed in *M* and *N* genes with 43% and 47%, respectively. However, the lowest value representative of the GC content in the second codon position (G+C2) was observed in the *E* gene (33%), and the maximum percentage in the *N* gene (50%). Whereas the minimum estimation of the GC composition in the third codon position (G+C3) was discovered in the *ORF1b* gene (24%) and the greatest proportion in both *M* and *N* genes (36%). Hence, the viruses used more AT% over GC%. The effective number of codons (ENC) values within the aforementioned genes ranged between 42.11 and 54.65 indicating a weakened codon bias. Similarly, the codon bias index (CBI) values varied between 0.36-0.40 and were in accordance with ENC results. Additionally, P values ≧ 0.05 were in favor of the null hypothesis assuming no codon bias (Table 1).

**Table 1.** Assessment of gene nucleotide composition and codon usage bias.

| Gene | ENC | CBI | SChi2 (P-value) | G+C2 | G+C3s | G+Cc |
| --- | --- | --- | --- | --- | --- | --- |
| **Wuhan_EPI_ISL_402123** |  |  |  |  |  |  |
| *ORF1a* | 44.49 | 0.40 | 0.36 (0.54) | 0.38 | 0.25 | 0.38 |
| *ORF1b* | 43.77 | 0.42 | 0.36 (0.54) | 0.38 | 0.24 | 0.37 |
| *S* | 44.38 | 0.43 | 0.35 (0.55) | 0.4 | 0.25 | 0.37 |
| *E* | 42.11 | 0.58 | 0.42 (0.51) | 0.33 | 0.32 | 0.38 |
| *M* | 54.65 | 0.38 | 0.21 (0.64) | 0.41 | 0.36 | 0.43 |
| *N* | 52.98 | 0.37 | 0.22 (0.65) | 0.5 | 0.36 | 0.47 |
| **Algeria_EPI_ISL_418241** |  |  |  |  |  |  |
| *ORF1a* | 44.45 | 0.40 | 0.36 (0.54) | 0.38 | 0.25 | 0.38 |
| *ORF1b* | 43.76 | 0.42 | 0.36 (0.54) | 0.38 | 0.24 | 0.37 |
| *S* | 44.36 | 0.43 | 0.35 (0.55) | 0.4 | 0.25 | 0.37 |
| *E* | 42.89 | 0.56 | 0.39 (0.53) | 0.33 | 0.32 | 0.38 |
| *M* | 54.65 | 0.38 | 0.21 (0.64) | 0.41 | 0.36 | 0.43 |
| *N* | 53.36 | 0.36 | 0.21 (0.64) | 0.5 | 0.36 | 0.47 |
| **Algeria_EPI_ISL_418242** |  |  |  |  |  |  |
| *ORF1a* | 44.45 | 0.40 | 0.36 (0.54) | 0.38 | 0.25 | 0.38 |
| *ORF1b* | 43.77 | 0.42 | 0.36 (0.54) | 0.38 | 0.24 | 0.37 |
| *S* | 44.36 | 0.43 | 0.35 (0.55) | 0.4 | 0.25 | 0.37 |
| *E* | 42.11 | 0.58 | 0.42 (0.51) | 0.33 | 0.32 | 0.38 |
| *M* | 54.65 | 0.38 | 0.21 (0.64) | 0.41 | 0.36 | 0.43 |
| *N* | 53.36 | 0.36 | 0.21 (0.64) | 0.5 | 0.36 | 0.47 |
| **Algeria_EPI_ISL_420037** |  |  |  |  |  |  |
| *ORF1a* | 44.44 | 0.40 | 0.36 (0.54) | 0.38 | 0.25 | 0.38 |
| *ORF1b* | 43.77 | 0.42 | 0.36 (0.54) | 0.38 | 0.24 | 0.37 |
| *S* | 44.36 | 0.43 | 0.35 (0.55) | 0.4 | 0.25 | 0.37 |
| *E* | 42.11 | 0.58 | 0.42 (0.51) | 0.33 | 0.32 | 0.38 |
| *M* | 54.65 | 0.38 | 0.21 (0.64) | 0.41 | 0.36 | 0.43 |
| *N* | 52.98 | 0.37 | 0.22 (0.65) | 0.5 | 0.36 | 0.47 |

Additionally, the relative synonymous codon usage (RSCU) analyses exhibited a similar pattern within both the reference sequence and the Algerian sequences with slight differences in the values. Overall, A3s and T3s were the most frequent nucleotides in the preferred codons and the least recurrent in the underrepresented ones among all investigated genes. In the *ORF1a* gene, among the twenty-six preferred codons, seventeen resulted with U while eight were A-ending and only one was G-ending. In consideration of the *ORF1b* gene, out of the twenty-seven preferred codons, sixteen were U-ending and eleven A-ending. Likewise, for the *S* gene, twenty-seven overrepresented codons englobed sixteen U-ending, nine A-ending and two G-ending. Similarly, sixteen overrepresented codons were observed in the *E* gene, eight counted as U-ending, five A-ending, two C-ending, and only one ended with a G. The *M* gene had twenty-three preferred codons comprising nine U-ending, eight A-ending five C-ending, and one G-ending. Lastly, regarding the *N* gene, twenty-five preferred codons were identified resulting in eleven U-ending, eight A-ending, five C-ending and one G-ending (Fig 4 and Tables S2-5).

**Fig 4.**
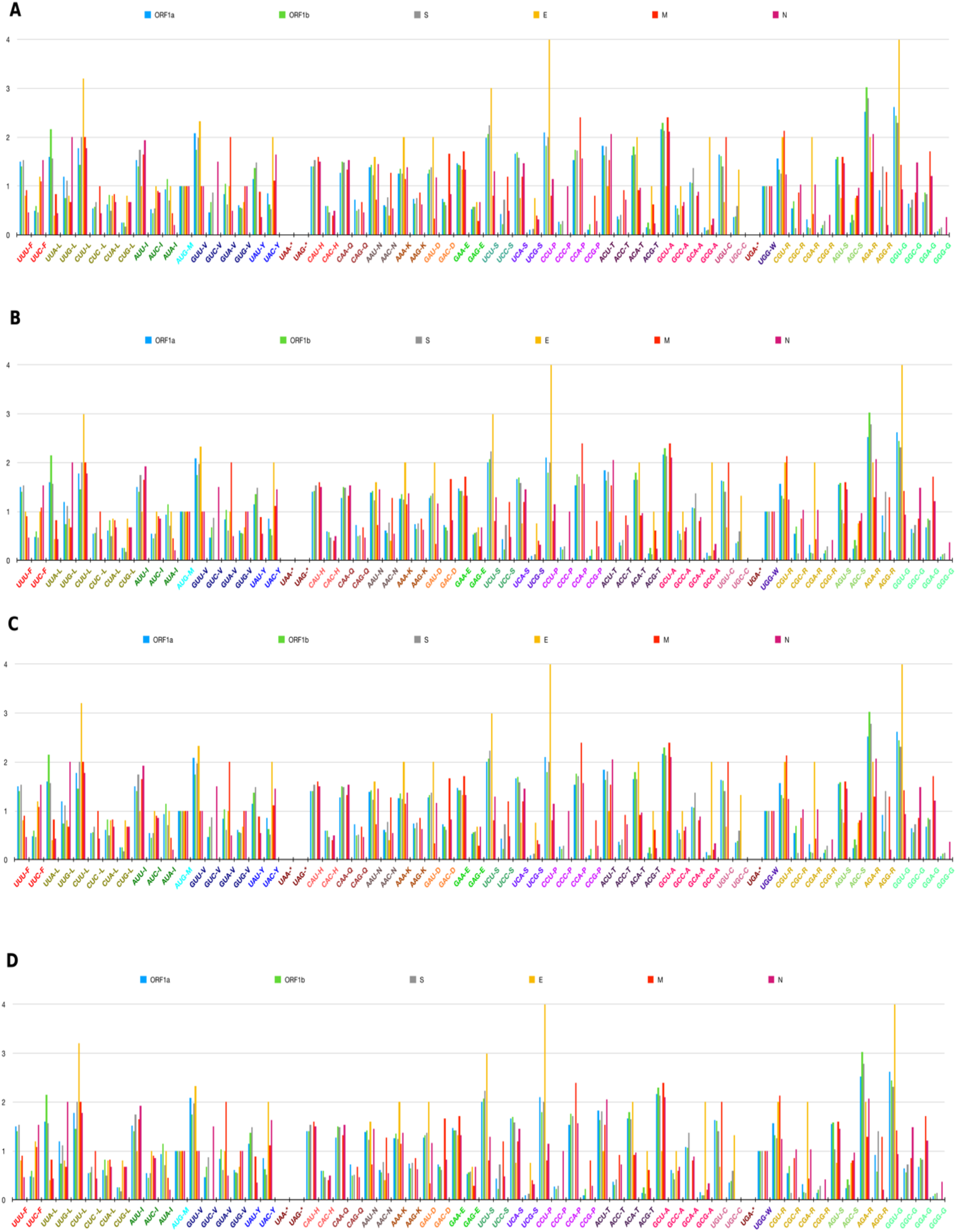
Relative synonymous codon usage (RSCU) analysis of the diverse codons of *ORF1a* (blue), *ORF1b* (green), *S* (grey), *E*(yellow), *M* (red), *N* (magenta) genes belonging to SARS-2-CoV genome. (**A, B, C, D**) represent the RSCU pattern of the reference sequence, Algeria_EPI_ISL_418241, Algeria_EPI_ISL_418242, and Algeria_EPI_ISL_420037 genomes, respectively.

Thereafter, pairwise Ka and Ks values were calculated representative of each gene. Only the Algerian and reference sequence data are depicted in Table S6. In the *ORF1a* gene, Ka/Ks ratios among each Algerian sequence and the reference sequence was < 1, indicating a purifying selection. Notably, when there is no variation (Ka=Ks=0), the (ω) ratio cannot be calculated (Algeria_EPI_ISL_418241 and Algeria_EPI_ISL_418242). Additionally, an infinite value was observed when Ks=0 and Ka>Ks (Algeria_EPI_ISL_418241 and Algeria_EPI_ISL_420037) which likely is in favor of a positive selection. In parallel, in consideration of the ORF*1b* gene, positive selection was observed in all sequence pairs except Algeria_EPI_ISL_418241 and Algeria_EPI_ISL_418242, in which the ratio was not calculable. Across the *S* gene, a positive selection was found when comparing the Algerian sequences to the reference sequence while the ka/ks ratio was not computable in the pairwise comparison regarding the comprehensive Algerian sequences. Similarly, in reference to the *E* gene, in which a positive selection was perceived when comparing Algeria_EPI_ISL_418241 to Algeria_EPI_ISL_418242, and Algeria_EPI_ISL_420037 to the reference sequence, whereas the Ka/Ks ratio was impossible to calculate in the remaining pairwise comparison. In consideration of the reliance to Ka and Ks values (ka=ks=0), a neutral evolution was observed in the *M* gene. However, the *N* gene exhibited a strong negative selection for all sequence pairs except in the cases of Algeria_EPI_ISL_418241 and Algeria_EPI_ISL_418242, including Algeria_EPI_ISL_418242 and the reference sequence where ka=ks=0.

Hereafter, based on Rambaut et al genome lineage attribution, the Algerian sequences fall within the lineage B1, together with the French sequences [6]. Additionally, both amino acid replacements and single nucleotide polymorphism (SNP) for each sequence compared to the reference sequence (NCBI RefSeq NC_045512) are summarized in Table 2. Strikingly, in comparing the Algerian sequences, whether to one another or to the French sequences, they displayed vast diversity at both the SNPs and amino acid replacement levels.

**Table 2.** SNPs and amino acid replacements of the Algerian sequences and the related French sequences.

| Sequence ID (Algeria) | SNP | Gene | AA replacement | Sequence ID (France) | SNP | Gene | AA replacement |
| --- | --- | --- | --- | --- | --- | --- | --- |
| **Algeria_EPI_I SL_420037** | C1059T<br>C3037T<br>**C5730T**<br>C10582T<br>C14408T<br>A23403G<br>G25563T | *nsp2*<br>*nsp3*<br>*nsp12*<br>*S*<br>*ORF3a* | T85I<br>**T1004I**<br>P323L<br>D614G<br>Q57H | EPI_ISL_437689 | C1059T<br>C3037T<br>C10582T<br>C14408T<br>A23403G<br>G25563T | *nsp2*<br>*nsp12*<br>*S*<br>*ORF3a* | T85I<br>P323L<br>D614G<br>Q57H |
| **Algeria_EPI_I SL_418241** | C1059T<br>C3037T<br>C10582T<br>C14408T<br>C18115T<br>A23403G<br>G25563T<br>**C25777T**<br>**C26461T**<br>**C29353T** | *nsp2*<br>*nsp12*<br>*nsp14*<br>*S*<br>*ORF3a*<br>*ORF3a*<br>*E* | T85I<br>P323L<br>**H26Y**<br>D614G<br>Q57H<br>**L129F**<br>**L73F** | EPI_ISL_414630 | C1059T<br>**C2003T**<br>C3037T<br>C10582T<br>C14408T<br>A23403G<br>G25563T<br>**C27059T** | *nsp2*<br>*nsp2*<br>*nsp12*<br>*S*<br>*ORF 3a* | T85I<br>**L400F**<br>P323L<br>D614G<br>Q57H |
| **Algeria_EPI_I SL_418242** | C1059T<br>C3037T<br>C10582T<br>C14408T<br>A23403G<br>G25563T<br>**C29353T** | *nsp2*<br>*nsp12*<br>*S*<br>*ORF3a* | T85I<br>P323L<br>D614G<br>Q57H | EPI_ISL_416498 | C1059T<br>C3037T<br>C10582T<br>C14408T<br>A23403G<br>**C23707T**<br>G25563T | *nsp2*<br>*nsp12*<br>*S*<br>*ORF 3a* | T85I<br>P323L<br>D614G<br>Q57H |

SNPs and Amino acid replacement proper to each sequence are indicated in bold, while the underlined SNPs are common to two sequences.

### Linkage disequilibrium

Recombination is characterized by a negative correlation between the distances among SNPs and the LD measures (D’ and r2). When plotting both the squared coefficient of correlation r2 and the normalized linkage disequilibrium coefficient D’ versus the distance between the polymorphic sites calculated in bases, no clear pattern of LD decay was observed (Fig 5A and 5B, respectively). Henceforth, a linear regression analysis to assess the aforementioned relationship was performed. The Pearson correlation coefficient between D’ and the distance was −0.01 and a coefficient of determination R2=0.0001, while between r2 versus the distance, the correlation coefficient was equal to −0.005 and R2=0.002. Furthermore, distances between sites in perfect LD (|D’|=1 and r2 >0.8) ranged between 1 and 26,409 bases, emphasizing the independence regarding the LD from the distance (Table S7). All results exclude recombination among SARS-CoV-2 studied genomes.

**Fig 5.**
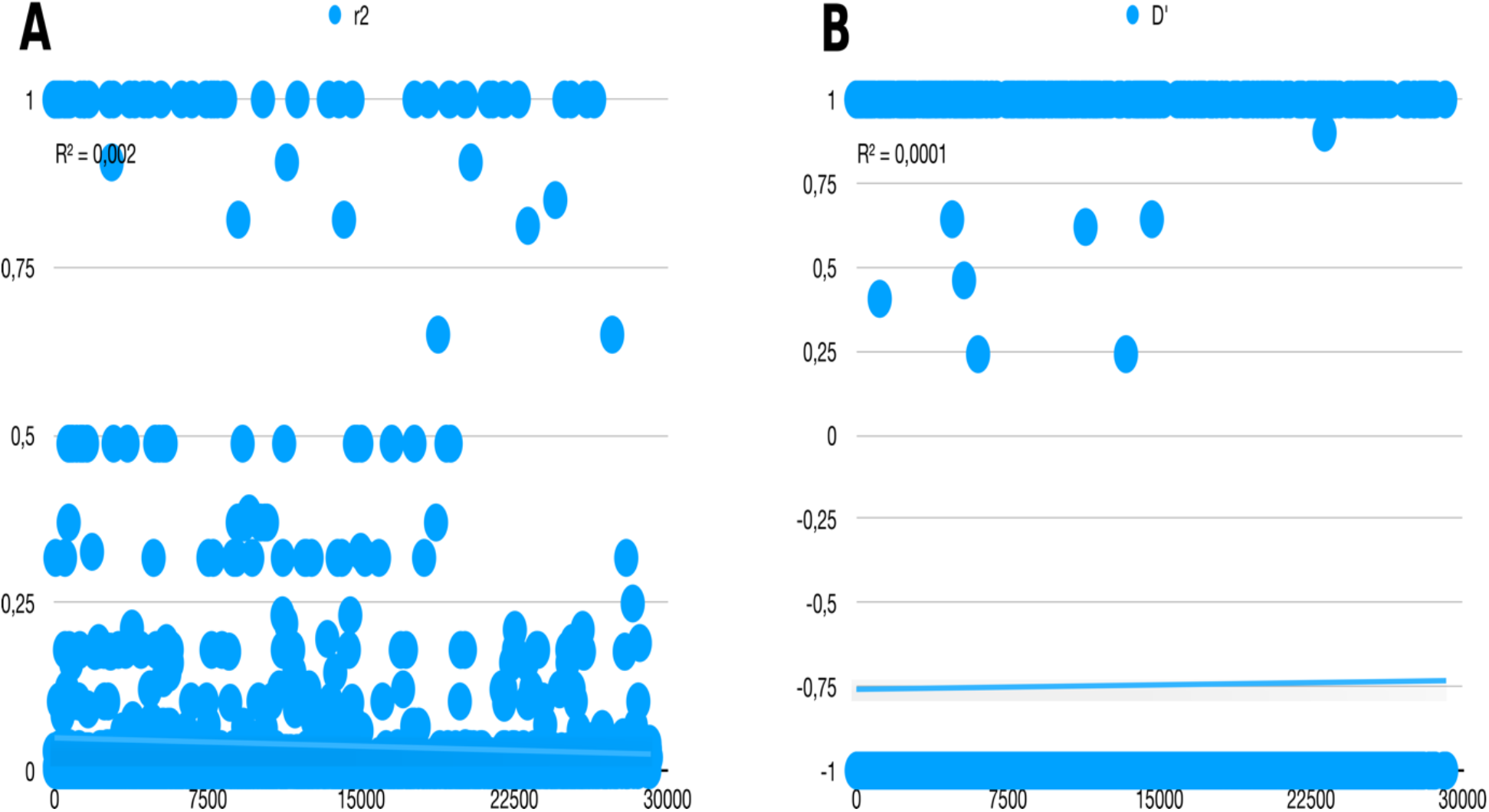
Linkage disequilibrium among polymorphic sites for forty-one SARS-CoV-2 complete genomes. (**A)** A linear regression analysis of the squared correlation coefficient r2 versus distance in bases. (**B)** A linear regression analysis of the normalized linkage disequilibrium coefficient D’ against distance in bases. The coefficient of determination R2 is indicated for both r2 and D’.

### Haplotype network analysis

Based on both RDP4 and DnaSP analysis, no recombination events were detected within the dataset. Thirty-six haplotypes were observed. The Algerian sequences clustered along with the French and the Belgian sequences. However, a median vector (small solid circle) was observed between France_EPI_ISL_414625, France_EPI_ISL_418218, Belgium_EPI_ISL_415159, and the Algerian sequences, in addition to Algeria_EPI_ISL_418241, Algeria_EPI_ISL_418242, and Algeria_EPI_ISL_410037 (Fig 6). Therefore, in lieu of the phylogenetic analysis, the results imply either missing data or unsampled sequences.

**Fig 6.**
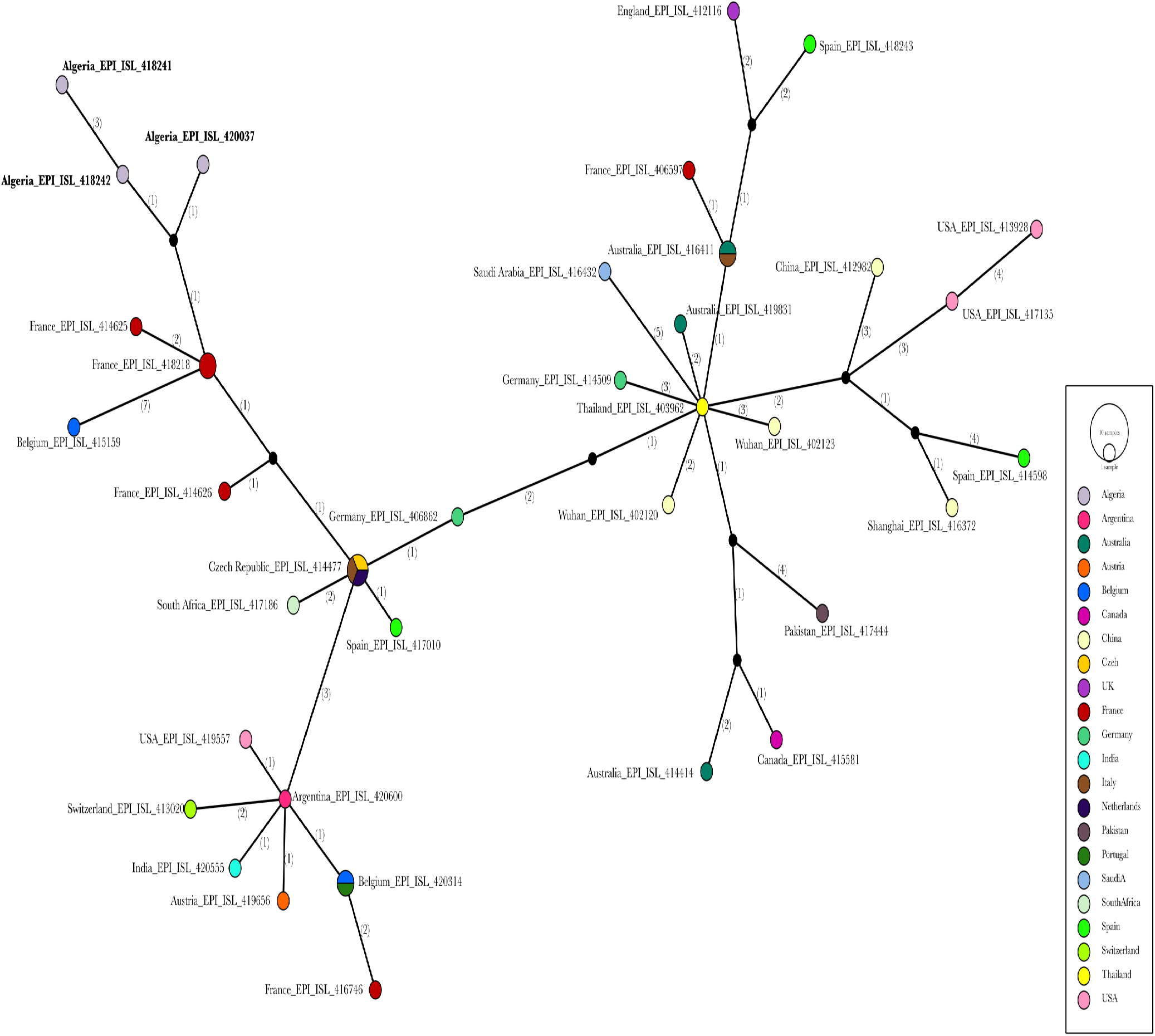
A haplotype network analysis representing forty-one SARS-CoV-2 genomes. The median joining algorithm with epsilon = 1 parameter was used in the network construction. Circles and diameters are proportional to the number of sequences. Mutation steps between haplotypes represent the numbers between brackets. All mutations were enclosed. The Algerian sequences are indicated in bold. Colors symbolize the different geographical sampling locations.

### Epidemiological analysis

The R2 values regarding the time series plot of the cumulative confirmed cases were 0.8, 0.9 and 0.5 for the exponential, linear, and logarithmic trend lines, respectively. Likewise, the same results were observed regarding recovery cases. Whereas the cumulative death cases exhibited the following R2 values: 0.6 for the exponential trend line, one for the linear, and 0.7 for the logarithmic model (Fig 7A). In parallel, the correlation coefficient calculated between the confirmed cases and the population density for each of the forty-eight Algerian cities was 0.69 (Fig 7B).

**Fig 7.**
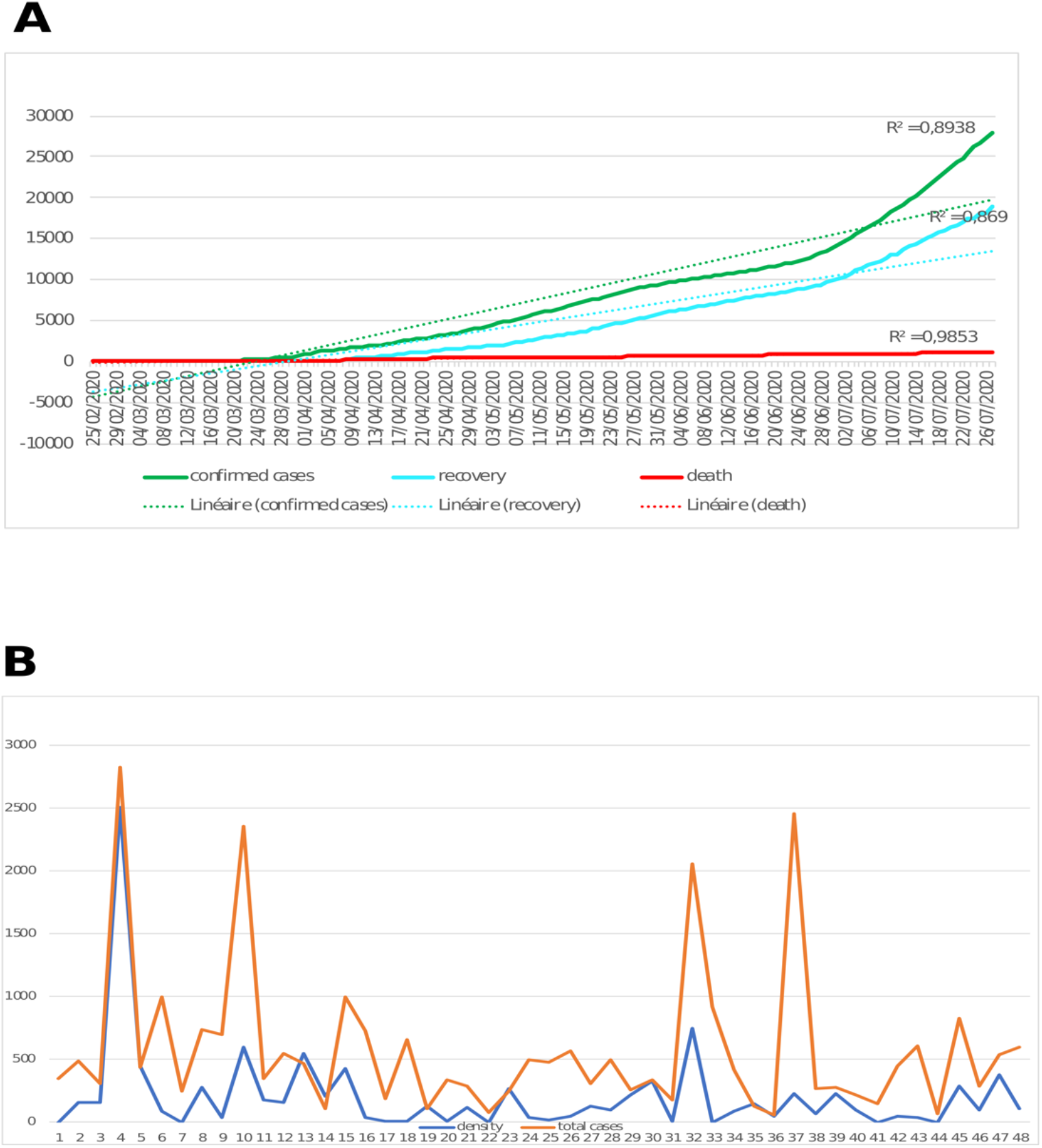
**(A)** Time series plot representing the COVID-19 confirmed cases (green), recovery (cyan), and death (red) depicting corresponding R2 values. **(B)** Correlation plot between the total confirmed cases and the population density for each Algerian city the numbers from 1 to 48 on the x-axis indicate each of the forty-eight Algerian cities.

## Discussion

To investigate the early spread of SARS-CoV-2 in Algeria, we performed a thorough analysis of forty-one SARS-CoV-2 complete genomes comprising all available Algerian sequences (n=3), in addition to sequences from France (n=6), Belgium (n=2), South Africa (n=1), Netherlands (n=1), Czech Republic (n=1), Spain (n=3), Italy (n=2), Argentina (n=1), India (n=1), Switzerland (n=1), Austria (n=1), USA (n=3), Portugal (n=1), Germany (n=2), Canada (n=1), Australia (n=3), Pakistan (n=1), Saudi Arabia (n=1), England (n=1), China (n=4) and Thailand (n=1).

Our estimations regarding the TMRCA of the SARS-CoV-2 global pandemic under a relaxed molecular clock with both the exponential growth and the skyline plot tree priors were in accordance with anterior studies performed on smaller or bigger datasets, since they are encompassed within their 95% HPD [7,8]. Similarly, the resultant TMRCA dates of the Algerian pandemic under the aforementioned tree priors, respectively, were estimated to 20/02/2020 [2020-02-13, 2020-02-27] and 21/02/2020 [2020-02-15, 2020-02-28]. These results are coherent, as the restriction measures in Algeria began from mid-March [5]. The evolutionary rate of the SARS-CoV-2 pandemic in the present study was equal to 0.001 substitution/site/year which is the exact substitution rate reported previously early in the pandemic [2,9].

In parallel, both the phylogenetic and the phylogeographic analyses supported the disease first originating in France and shortly thereafter migrating to Algeria, (PP=0.9 and BF=42.04) as previously demonstrated through contact tracing [5]. Interestingly, areas in which missing data was clearly obvious was made apparent through phylogenetic reconstruction. Moreover, the phylogeographic analysis highlighted the route of spread regarding the pandemic originating from the epicenter, China, throughout Asia, then onto Europe and finally to Africa. Despite the small size of the dataset, it is in accordance with the large SARS-CoV-2 phylogeographic analysis performed in Nextstrain [10].

Additionally, the phylodynamic analyses revealed a growth factor of 0.06/day (23.9/year) resulting in a doubling time of 11.6 days which is aligned with similar estimates performed early in the pandemic [11,12]. Moreover, the Bayesian skyline plot revealed a logistic growth regarding the effective population size in the early pandemic (from 24 December up through the 8 March). This may likely be justified, since, at the beginning of this time-lapse, only a few sequences were uploaded to the GISAID database. Moreover, at this stage, the disease qualified as an Wuhan outbreak, and not many genomes were sequenced worldwide [13,14]. From mid-January up through mid-February, more sequences were available from China and only minute sequences from other countries. Inexplicably, very few genomes were accessible, whether from China nor from the rest of the world in the waning two weeks of February. This period represented the silent global spread of SARS-CoV-2 including Algeria, since asymptomatic individuals were not considered regarding sampling [15].

Overall, The Algerian SARS-CoV-2 sequences displayed a lower genetic diversity with genomes from different geographic origins revealing little differentiation. The same results were observed in a recent study among 18,514 sequences including the Algerian genomes [16]. Furthermore, when comparing the Algerian sequences to the reference sequence, the codon bias analysis revealed very similar results amid the forgoing genomes. Low GC values were observed in all investigated genes (ORF1a, ORF1b, M, N, S and E), whether in the global composition or both the second and the third codon positions. Moreover, ENC values in different genes ranged between 42.11 and 54.65, whereas CBI estimates fluctuated between 0.37 and 0.58, thus, indicating the potential bias regarding slight codon usage. These results are aligned with anterior studies performed on SARS-CoV in the same context implying a low bias regarding codon usage among RNA viruses with little variation between genes due to their high mutation rate [17]. From another viewpoint, the RSCU pattern across the Algerian and the reference genomes were very similar with slight differences regarding the values in all genes, in addition to a preference for U-ending codons in the overrepresented codons. Once more, as per anterior reported studies regarding the SARS-CoV-2 codon usage, the pattern is more or less conserved independently from the geographical provenance [18,19]. Globally, the ω ratio across all genes was often times either impossible to calculate (ka=ks=0) or had extreme values (∞ when ks=0 and 0 when ka=0), of which is due to insufficient nucleotide variations characteristic in the early phase of the pandemic [12,20]. Moreover, the small data panel of the Algerian sequences (n=3) is a drawback regarding the comprehension of the selection pressure pattern within Algeria, of which, is an important implication in diagnostics, therapy, and vaccine development. In view of the fact in which Algeria was under complete travel restrictions since the 15 March, the number of cases kept increasing, indicating local transmissions, thus these local viral variants may potentially represent a distinct strain [21].

Complementary to this, several common non-synonymous mutations were detected among the Algerian sequences and the related French sequences. This included T85I in the nsp2 gene, P423L in the nsp12 gene, D614G in the S gene, and Q57H in the ORF3a gene. Notably, these mutations were detected within eighty-four countries and thus considered as positively selected [22–24]. Interestingly, unique non-synonymous mutations were detected. To cite and example, T1004I replacement in the nsp3 gene was detected in the sequence Algeria_EPI_ISL_420037. This mutation was spotted as a unique mutation in the USA in the early stages of the pandemic, sequences from 19 January to 15 April, and was not reported elsewhere. Since we know three Algerian sequences were derived from locals without any travel history, we can conclude the individual who contaminated Algeria_EPI_ISL_420037 had either a travel history to the USA or was in close contact with an individual who introduced the disease to France originating from the USA [25]. Strikingly, in Algeria, _EPI_ISL_418241, H26Y, L129F, and L73F mutations were identified in *nsp14, ORF3a* and *E* genes, respectively. According to a recent study regarding SARS-CoV-2 mutations, the H26Y mutation was defined as a low-frequency non-synonymous mutation of a second phase infection (sequences between 18 April – 17 May), nevertheless, it was detected in our sequence sampled on the 2 March. Moreover, the L129F non-synonymous mutation in the ORF3a gene was proven to be characteristic only in reference to our Algerian sequence among 2,782 analyzed sequences and is considered under negative selection [26]. Additionally, L73F amino acid replacement was revealed in the E gene, in which this mutation was reported in Australia and consequently, is the alteration of the DLLV motif (change to DFLV). Distinctly, it may delay Tight Junction formation and therefore may hypothetically affect viral replication and/or infectivity [27].

Notably, only the sequence Algeria_EPI_ISL_418242 exhibited an identical mutational pattern regarding the protein level when compared with the related French sequences (identical to EPI_ISL_437689 and EPI_ISL_416498). Furthermore, the haplotype network analysis was in accordance with the aforementioned results and displayed a median vector between both the Algerian and the French sequences, in addition to the two Algerian sequences Algeria_EPI_ISL_420037 and Algeria_EPI_ISL_418242, indicating unsampled data. This conformed to a French study on tracing the introduction of SARS-CoV-2 to France, which included all the French sequences and the Algerian sequences. We observed how Algeria_EPI_ISL_420037 didn’t cluster with the two other Algerian sequences. Moreover, the branch length which was proportional to nucleotide substitution indicated clearly unsampled data between Algeria_EPI_ISL_418242 and Algeria_EPI_ISL_418241 [28]. Unsurprisingly, the three Algerian sequences were classified in the B.1 lineage which is the common circulating virus in both Europe (France) and North America (USA) [29].

Among these findings, our results supporting the absence of recombination among SARS-CoV-2 in the early phase of the pandemic and are in accordance with a previous study performed on 37566 SARS-CoV-2 sequences [30]. However, VanInsberghe and colleagues were able to detect five putative recombinant genomes among 47,390 unique sequences, despite the low recombination percentage (0.007%), thus, providing evidence regarding the increase of recombination possibilities in parallel to the pandemic expansion [31].

In the present study, we demonstrated the evolution representing the Algerian pandemic in a consistent manner reflecting the effectiveness of varied implemented measures. Moreover, the moderate correlation between the number of SARS-CoV-2 confirmed cases and the population density in each Algerian city implies the spread of the virus is primarily dependent on the awareness of the community and the respectful compliance regarding social distancing, as lower infection cases in relatively high population density cities were observed and vice-versa. To cite an illustration, Adrar, a city located in the southern portion of Algeria, has a population density of 1.03 habitant/km2 and as of 27 July, the number of confirmed cases is 350, whereasas, in Jijel, situated in the Northern area of Algeria, the number of confirmed cases is 246 cases for 265.5 habitant/km2. This is complementary to an epidemiological study conducted to assess the mitigation measures implemented in Algeria in the early SARS-CoV-2 pandemic (dated 26 April), which demonstrated the efficiency based on the basic reproduction number R0 before and after the implementation of the preventive strategy [5].

Overall, we explored the evolution, genetic, and the epidemiological aspects regarding the Algerian SARS-CoV-2 during the early phase of the pandemic, aptly demonstrating the introduction of the disease and the heterogeneity of the genomes. Additionally, our research findings revealed a unique amino-acid substitution and a closeness to USA sequences by characterizing the mutational pattern. Statistically, we assessed the effectiveness regarding the mitigation majors implemented against the SARS-CoV-2 pandemic. Admittedly, the main drawback regarding our study was the size of the Algerian data panel. Thus, we emphasized the importance of massive sampling and sequencing in disease comprehension and increased efforts regarding drug and vaccine development.

## Materials and methods

### Sequence selection and maximum likelihood phylogeny

Forty-one sequences originating from the current SARS-CoV-2 pandemic including the three Algerian genomes were retrieved from the GISAID database [13,32]. All data samples were acquired and prepared prior to 8 March 2020, which is the latest collection date in reference to the Algerian sample (Table S8). Therefore, sequences were aligned by MAFFT using default parameters [33]. Subsequently, a maximum likelihood phylogenetic tree was implemented in IQTREE webserver, under the HYK+G4 substitution model, with ultrafast bootstrapping [34, 35].

### Temporal signal assessment, time-calibrated phylogeny reconstruction, and Phylogeographic analysis in discrete space

To effectively assess the clock-likeliness regarding the data, the aforementioned maximum likelihood resultant tree was used as an input file in TempEst [36]. A regression analysis of root-to-tip genetic distance against sampling time demonstrated a strong positive correlation/correlation coefficient= 0.76 while a moderate association was observed /R^2^=0.57, indicating the suitability of the dataset for a phylogenetic molecular clock analysis. Subsequently, the tip dated phylogenetic tree was generated using the Beast v1.10.4 package and employing the HKY+G4 substitution model. Moreover, both strict and lognormal uncorrelated relaxed clock models were tested [37, 38]. In consideration of population size and growth, the parametric coalescent exponential growth model assuming an exponential increase in the population and the non-parametric Skyline plot model supposing different effective population sizes for each coalescent interval was applied as priors [39, 40]. To estimate both the MRCA of the Algerian sequences including the time stretching back to the Algeria MRCA /TMRCA, a taxon set comprising the Algerian sequences was created. The MCMC chains were operational for 100 million generations and sampled every 10,000 generations, with 10% discarded as burn-in. Subsequently, the effective sampling sizes /ESS>200 were examined using TRACER v1.6.0 [41]. In parallel, the date of the most recent common ancestor/MRCA regarding the pandemic in addition to the evolutionary rate was estimated. Furthermore, the Maximum clade credibility tree/MCC was annotated by means of TreeAnnotator v1.10.4 and visualized in FigTree v1.4.4 [21]. Additionally, a phylogeographic analysis was performed. The samples’ spatial data/location of isolation was used to infer the geographical spreading patterns of the virus by combining the Bayesian stochastic search variable selection/BSSVS with a standard symmetric substitution implemented using Beast v1.10.4. [42]. Thereafter, the MCC tree was loaded in SpreaD3 v0.9.7 to visualize and analyze the transmission routes, and the log file to calculate the Bayes Factor/BF [43].

### Genome investigations

MEGA X was first used to calculate the pairwise genetic distances employing the p distance parameter [44]. Subsequently, the DnaSP v6.12.03 package was applied in reference to further genome exploration, beginning with polymorphic region assessment. Furthermore, the presence of insertions or deletions (InDels) was estimated based on the diallelic model. Thereafter, for each of the following genes: *ORF1a, ORF1b, S, E, M, N*, several indices of codon usage bias were calculated, comprising the frequency of the nucleotides G+C at the second and third position (G+C2, G+C3s) and the GC contents in the coding region (G+Cc). the effective number of codons (ENC) used to measure the degree of the codon usage bias. The ENC values range from twenty (when only one codon is used for each amino acid) to sixty-one (when all synonymous codons are used for each corresponding amino acid). Generally, a significant codon usage bias in a coding region is indicated by an ENC value less than or equal to 35 [45]. Codon Bias Index (CBI) was used to estimate the codon usage bias based on the fraction of preferred codons used in a gene, and its values range from zero (uniform use of synonymous codons) to one (only preferred codons are used) [46]. Additionally, the relative synonymous codon usage (RSCU) evaluating the ratio of the observed frequency to the expected one for each codon assuming all codons for a particular amino acid are used equally. An RSCU value <1.0 represents a negative codon usage bias while RSCU >1.0 implies a positive bias, and a value equal to one is interpreted as no bias. To assess the significance of the codon usage bias, a scaled chi-square test (SChi2) was performed. Only the results of the SARS-CoV-2 reference sequence and the Algerian sequences were reported. Likewise, to assess the selective pressure at the protein level, (ω) the ratio of the non-synonymous mutation rate (Ka) to the synonymous mutations (Ks) was evaluated within each gene for each sequence pair according to Nei and Gojobori [30,47]. When several nonsynonymous mutations which promote changes with physiochemically different amino acids occur, they show a tendency to be deleterious to the protein, and thus they are improbable to become fixed in the population leading to a negative selection resulting in Ka<Ks (ω<1). Contrariwise, when advantageous nonsynonymous substitutions strike, they are likely to become fixed in the population, and thus amino acid changes in the protein are enhanced (ω>1). Thereafter, linkage disequilibrium between nucleotide variants was estimated. All polymorphic sites were analyzed to test the recombination among SARS-CoV-2 genomes within the dataset. The linkage disequilibrium (LD) decay signal which is a recombination characteristic was examined. When recombination occurs, it will disrupt linkage between SNPs, the more distant SNPs will be affected faster and the significance of the results was based on Both the two-tailed Fisher’s exact test and the chi-square test with Bonferroni correction (*, P < 0.05; **, P < 0.01; ***, P < 0.001) [48, 49].

Lastly, we subjugated the Algerian sequences and three different French sequences selected based on an ulterior study conducted by Gámbaro and colleagues to the CoV-GLUE webserver to highlight the differences among them and the related French sequences and further to reveal the lineage of the Algerian genomes and the amino-acid variations [6, 28, 50].

### Haplotype network analysis

Prior to haplotype network analysis, RDP 4 software was used for recombination detection, thus, the median-joining network method implemented in POPART software was employed with default setting/epsilon=0 [51–53].

### Epidemiological analysis and preventive measures assessment

In summary, to draft an overview encapsulating the evolution of the pandemic in Algeria, the cumulative number of the infected recovered and death cases were collected from the Johns Hopkins University Center for Systems Science and Engineering (27 July 2020) [54]. Subsequently, the linear, exponential, and the logarithmic trend lines were compared, and the best model was chosen based on the R2 values. Furthermore, the cumulative confirmed cases for each of the forty-eight Algerian cities were collected from the official Algerian ministry of health website [55]. Additionally, the population density data regarding all Algerian cities was retrieved from the Wikipedia website [56]. Thereafter, the correlation coefficient was calculated between the density and the number of confirmed cases.

## Data Availability

All data are available within the manuscript and the supplimentary material file.

## Acknowledgments

We gratefully express our appreciation and wish to thank the authors, originating and submitting laboratories of the sequences from GISAID’s EpiFlu(tm) Database, on which this research is based. We acknowledge Dr. Vincent Enouf for performing the NGS sequencing for our Algerian samples.

## Author contributions

Conceptualization: SZ, BS, and GK. Performed the analyses: SZ, RH and BZ. Sample collection: FD. Original draft preparation: SZ and GK. Supervision: FJ

## Supporting information

**S1 Table. Estimates of Evolutionary Divergence over Sequence Pairs between Groups**. The number of base differences per site from averaging over all sequence pairs between groups is shown. Standard error estimate/s are shown between brackets and were obtained by a bootstrap procedure /1000 replicate.

**S2 Table. The relative synonymous codon usage (RSCU) pattern for the reference sequence (Wuhan_EPI_ISL_402123)**. Over expressed codons are highlighted in cyan whereas the under expressed codons are highlighted in yellow.

**S3 Table. The relative synonymous codon usage (RSCU) pattern for Algeria_EPI_ISL_418241**. Over expressed codons are highlighted in cyan whereas the under expressed codons are highlighted in yellow.

**S4 Table. The relative synonymous codon usage (RSCU) pattern for Algeria_EPI_ISL_418242**. Over expressed codons are highlighted in cyan whereas the under expressed codons are highlighted in yellow.

**S5 Table. The relative synonymous codon usage (RSCU) pattern for Algeria_EPI_ISL_420037**. Over expressed codons are highlighted in cyan whereas the under expressed codons are highlighted in yellow.

**S6 Table. Synonymous and non-synonymous mutation rates estimations for the Algerian sequences and the reference sequence**. ω = Ka/Ks ratio, - impossible to calculate (Ka=Ks=0), Algeria 1: Algeria_EPI_ISL_418241, Algeria 2: Algeria_EPI_ISL_418242, Algeria 3: Algeria_EPI_ISL_420037, reference: Wuhan_EPI_ISL_402123.

**S7 Table. Pairwise linkage disequilibrium among polymorphic sites for 41 SARS-CoV-2 genomes. Dist**. distances in bases, **r**. Correlation coefficient, **r2**. squared coefficient of correlation, **D**. linkage disequilibrium coefficient, **D’**. the normalized linkage disequilibrium coefficient. **\*** P<0.05; **\*\*** P<0.01; **\*\*\*** P<0.001. **B** indicates Bonferroni correction. Sites in perfect LD are highlighted in yellow.

**S8 Table. Details of sequences used in this study**. Accession ID, sequences names, date of collection and country of provenance.

